# Neighborhood-level Racial/Ethnic and Economic Inequities in COVID-19 Burden Within Urban Areas in the US and Canada

**DOI:** 10.1101/2020.12.07.20241018

**Authors:** Sudipta Saha, Justin M. Feldman

## Abstract

The COVID-19 pandemic exhibits stark social inequities in infection and mortality outcomes. We investigated neighborhood-level inequities across cities in the US and Canada for COVID-19 cumulative case rates (46 cities), death rates (12 cities), testing rates and test positivity (12 cities), using measures that characterize social gradients by race/ethnicity, socioeconomic composition, or both jointly. We found consistent evidence of social gradients for case, death and positivity rates, with the most privileged neighborhoods having the lowest rates, but no meaningful variation in the magnitude of inequities between cities. Gradients were not apparent in testing rates, suggesting inadequate testing in the most deprived neighborhoods. Health agencies should monitor and compare inequities as part of their COVID-19 reporting practices and to guide pandemic response efforts.

**HIGHLIGHTS:**

- Within urban regions with available data in the US and Canada, there were strong social gradients for case, death and positivity rates
- The most racially and/or economically privileged neighborhoods had the lowest rates
- Social gradients were similar for neighborhood-level measures of racial/ethnic composition, income, racialized economic segregation, and racialized occupational segregation
- Testing rates did not show consistent social gradients, which suggests that the most deprived neighborhoods have inadequate access to testing relative to their higher disease burden

## INTRODUCTION

Similar to numerous other health outcomes, the impact of Coronavirus Disease 2019 (COVID-19) has been marked by racial/ethnic and class inequities. In the US, the CDC currently reports racial inequities in COVID-19 cases, hospitalizations and deaths. According to this data, Non-Hispanic Black / African American, Hispanic or Latino, and American Indian / Alaska Native people are around three times as likely to have been diagnosed with COVID-19, around five times more likely to be hospitalized, and up to two times more likely to die compared to white people (CDC, 2020). However, the quality and completeness of disaggregated data has varied from state to state, and approximately 20% of confirmed COVID-19 cases are missing race/ethnicity data in the US (The COVID Tracking Project, 2020). Individual-level socioeconomic data have been largely absent in reporting. In neighboring Canada, which also has had long-standing racial and class inequities in health outcomes, data stratified by race and socioeconomic position has been largely absent (Shahidi et al., 2020; Tjepkema et al., 2013). Notable exceptions include some local public health units, like Toronto, which has reported that rates of confirmed cases among visible minorities (especially Black people) are higher relative to those of white people, and has also reported a strong gradient by household income (City of Toronto, 2020).

In the absence of individual-level socioeconomic data, area-based measures can highlight gradients in socioeconomic inequities for COVID-19 outcomes (Gordon, 2003; Krieger et al., 2003, 2002). Moreover, neighborhood-level analyses can capture contextual factors that are emergent at higher levels than the individual (e.g. concentrated economic segregation within urban areas). While urban areas have been heavily impacted by COVID-19, to our knowledge there has not been a comprehensive assessment of COVID-19 inequities within cities in Northern America. Studies to date have focused on particular cities individually, especially those that emerged as the first hotspots in the US (Benitez et al., 2020; Chen and Krieger, 2020; Cordes and Castro, 2020; Lieberman-Cribbin et al., 2020, pp. 1-; Maroko et al., 2020; van Holm et al., 2020). In the past few months, many local jurisdictions have started disseminating case, death and testing data at postal-code level or for other small geographic areas. We collected reported data from major urban areas in the United States and Canada and investigated neighborhood-level inequities.

Our analysis is guided by an understanding of racial capitalism as a fundamental cause of COVID-19 - a framing discussed in recent papers by William Pirtle, and McClure et al (McClure et al., 2020; Pirtle, 2020). Theoretically developed by Cedric Robinson, “racial capitalism” refers to a system where “as a material force, … racialism … inevitably permeate[s] the social structures emergent from capitalism” (Robinson, 1983). In the context of COVID-19, racial capitalism manifests as increased risks for Black and Hispanic people due to segregation and inequities in occupation, housing conditions, access to healthcare, and also victimization by mass incarceration.

Under this theoretical framework, we investigate neighborhood-level inequities along gradients of income, racism, racialized economic segregation, and racialized occupational segregation. We measure racialized economic segregation using Index of Concentration at the Extremes (ICE) for race and income, which has been used previously for a number of different health outcomes, as well as for COVID-19 in some settings (Chen and Krieger, 2020; Feldman et al., 2015; Krieger et al., 2015). We also introduce a new measure - ICE for race and occupational class – to capture racialized segregation of high and low risk occupations.

## METHODS

### Data source

We visited health department websites for the 69 urban counties (as defined by the National Center for Health Statistics) in the United States to identify counties for which data were available at sub-county and sub-city geographic levels (National Center for Health Statistics, 2019). When county-level data were not available, we determined whether it was available from the health department of the largest city in that county. We additionally queried the health department websites for the four largest cities in Canada. In most cases, data was reported by Zip Code Tabulation Area (ZCTA), while for Washington DC, Toronto ON and Montréal QC, data was reported by municipally defined neighborhood boundaries. Jurisdictions that shared a border were merged to create combined urban regions, since patterns of urban spatial segregation likely traverse across boundaries of neighboring urban counties (e.g. New York City counties/boroughs). These merged regions are hereafter referred to as “cities”, and nested smaller areas as “neighborhoods”, for simplicity. Details of data availability by location is outlined in **Supplementary Table 1**.

### Measures

Our primary outcome of interest was the rate of cumulative COVID-19 cases in each neighborhood, while our secondary outcomes of interest were rates of cumulative deaths, tests and test positivity, where data was available.

We created quintiles *within* each city for each variable of interest, with the most privileged quintile being set as the reference category. Variables reported here are: median household income, percentage of non-white residents, ICE for race and income, and ICE for race and class. ICE is calculated as: 

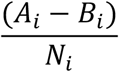

Where *A*_*i*_ represents the number of people in the most privileged group, *B*_*i*_ the number of people in the most deprived group and *N*_*i*_ the total population in neighborhood *i*. The index ranges from −1 to 1, corresponding to two extremes of spatially concentrated deprivation and privilege respectively (Krieger et al., 2015; Massey, 2001). For race + income, the most privileged group was defined as non-Hispanic white people who have an income greater than $100,000, and the most deprived group was defined as non-Hispanic Black people who have an income less than $25,000. For race + class, the most privileged group was defined as white workers in “management, professional and related occupations”, which are largely amenable to work-from-home arrangements. The most deprived group included Black workers in “service”, “natural resources, construction, and maintenance operations”, and “production, transportation, and material moving” occupations. These were generally considered essential, yet underpaid and high-risk for SARS-CoV-2 exposure. The groups in neither extreme would include workers of other racial groups, and workers in “sales and office” occupations. These include a mix of jobs that could be performed from home, as well as some frontline retail work. However, since data is only available for coarse categories of occupation by race by ZCTA, we considered this group to be at neither extreme. ICE measures were only available for US cities due to lack of readily available data by race and income or occupation in the Canadian Census. Canadian cities were dropped when ICE measures were used.

For the United States, data for all variables was obtained from American Community Survey (2014-18) at the ZCTA level, or CT level to be aggregated into municipally defined neighborhoods. For Canada, data was obtained from long-form Census variables at the Census dissemination area (CDA)-level and aggregated up to municipally defined neighborhoods. Where CDA’s crossed boundaries, numbers were reallocated weighted by spatial overlap.

### Statistical Analysis

We present rate ratios of cumulative cases, deaths, and tests, and test positivity by quintile. Inequities across cities were modelled using separate multi-level Poisson regression for each outcome, and each social measure. We included random intercepts for each city, and observation (neighborhood)-level random intercepts to account for overdispersion. For each model, we included all available cities with the available outcome data. To ensure that our general conclusions were robust, we repeated the analysis on only the subset of cities that had data for multiple outcomes present. Finally, to determine if inequities varied between cities, we separately fit models with random intercepts and slopes for quintiles as a continuous variable, and assessed estimates of the random slopes variance.

We present detailed city-specific plots and outcomes, for the reader to explore at: https://ssaha.shinyapps.io/COVID_cities_inequities/. The dataset can also be accessed at: https://github.com/saha-sudipta/COVID_Cities_Disparities

## Results

Fifty-eight of 69 US urban counties (or the largest city in an urban county) had case data available at smaller geographic units. Two of the four largest cities in Canada had data available. In total, these represented a population of 85 million, with 1.8 million cumulative cases. At the time of data collection, this represented more than a quarter of all reported cases in the US and Canada. After merging adjacent jurisdictions, there were 46 “cities” in total. Of the 46, 12 reported COVID-19 deaths by neighborhood, and 12 reported tests by neighborhood (**Table 1**).

**Table 1:** Summary statistics of populations and outcomes

|  | Cases | Deaths | Tests |
| --- | --- | --- | --- |
| **Population included** | 84961152 | 27748074 | 24608418 |
| **Number of urban regions included** | 46 | 12 | 12 |
| **Total cumulative outcomes** | 1806258 | 30420 | 6406629<br>Positive: 525657 |
| **Total cumulative outcome rate per 100,000** | 2125.98 | 109.62 | 26034<br>Positivity: 8.20% |
| **Median cumulative outcome rate at city-level [IQR]** | 1978.03<br>[1389.00, 2501.73] | 32.77<br>[21.04, 127.86] | 24151<br>[18838, 29809]<br>Positivity:<br>6.70%<br>[5.97%, 9.95%] |
| **Median cumulative outcome rate at neighborhood-level [IQR]** | 1736.75<br>[970.55, 2648.73] | 37.82<br>[11.41, 130.25] | 18147<br>[25111, 32150]<br>Positivity:<br>6.90%<br>[4.61%, 10.01%] |

At the city-level, the cumulative case rates ranged from 265.9 per 100,000 in Multnomah County, Oregon to 5977 per 100,000 in Miami-Dade County, Florida (median 1978 per 100,000). Among the 12 cities with test data available, the cumulative testing rate ranged from 12.65 per 100 people in Alleghany County, Pennsylvania, to 43.17 per 100 people in New Orleans Parish, Louisiana (median 24.15 per 100 people). The positivity rate ranged from 4.45% in Alameda County, California to 10.6 % in Milwaukee, Wisconsin (median 7%). Among the 12 regions with death data, the cumulative death rate ranged from 3.21 per 100,000 people in San Francisco - Santa Clara, California, to 226 per 100,000 people in New York City (median 32.77 per 100,000) (**Supplementary Table 2**).

Neighborhood-level sociodemographic measures were correlated with one another to varying degrees. Quintiles of median household income had correlations of −0.53, 0.72, and 0.86 with percentage non-white population, ICE race + class, and ICE race + income respectively. Percentage non-white had correlations of −0.67 with both ICE race + income and ICE race + class. The two ICE measures had a correlation of 0.86.

With some exceptions, the most privileged quintile had lower cumulative case, death and positivity rates, compared to the most deprived quintile in each city. Testing rates did not show consistent patterns.

In our modeled estimates of inequities across urban regions, there were strong and monotonic social gradients in cumulative case incidence, deaths and positivity across quintiles of all sociodemographic measures. The choice of neighborhood measure did not substantially affect the magnitude of inequities on the rate ratio scales. The rate ratio for cumulative case incidence, comparing the most deprived quintile to the most privileged quintile, was significant using all measures, with ICE race + income having the highest effect size (RR: 2.08; 95% CI: 1.95, 2.21), and ICE race + class having the lowest (RR: 1.88; 95% CI: 1.77, 2.01). For test positivity, the highest effect size was for ICE race + class measure (RR: 2.41; 95% CI: 2.23, 2.61), and the lowest for median household income (RR: 2.03; 95% CI: 1.86, 2.22). The magnitude of inequities in mortality were higher than for other outcomes, with the highest rate ratio present from the ICE race + income measure (RR: 3.59; 95% CI: 3.05, 4.24), and the lowest for percentage non-white (RR: 2.77; 95% CI: 2.29, 3.36). Greater inequities for mortality persisted when the analysis was limited to cities that had both case and death data (**Supplementary Table 3**).

There were no clear and persistent socio-economic gradients for rates of testing, although for ICE race + class there was a trend for fewer tests in deprived neighborhoods, with the most deprived quintile having the lowest rate compared to the most privileged (RR: 0.77; 95% CI: 0.70, 0.85) **(Figure 1, Table 2)**. The discordance between case and testing rates persisted when the analysis was limited to cities with both case and testing data (**Supplementary Table 4**).

**Table 2:**
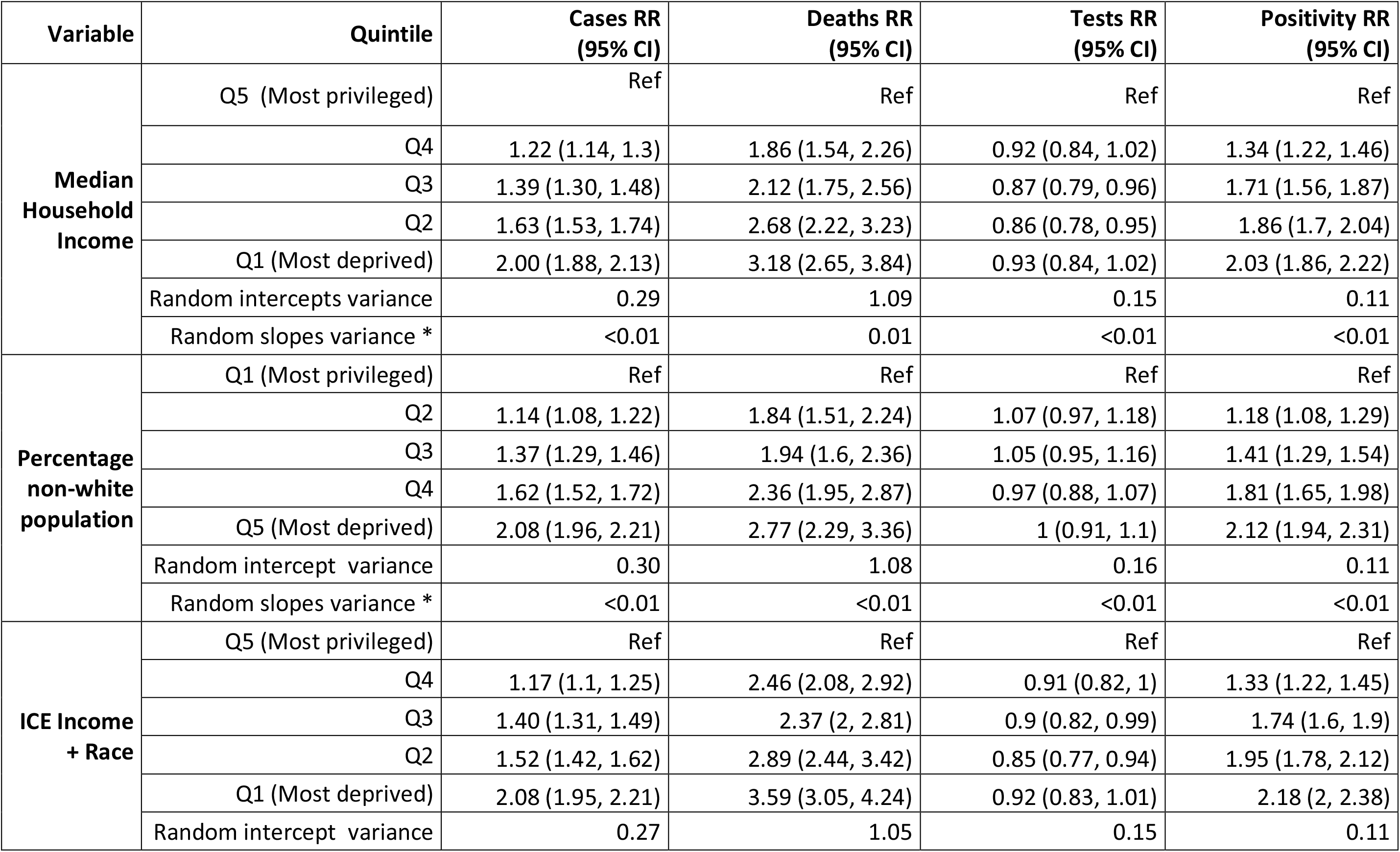
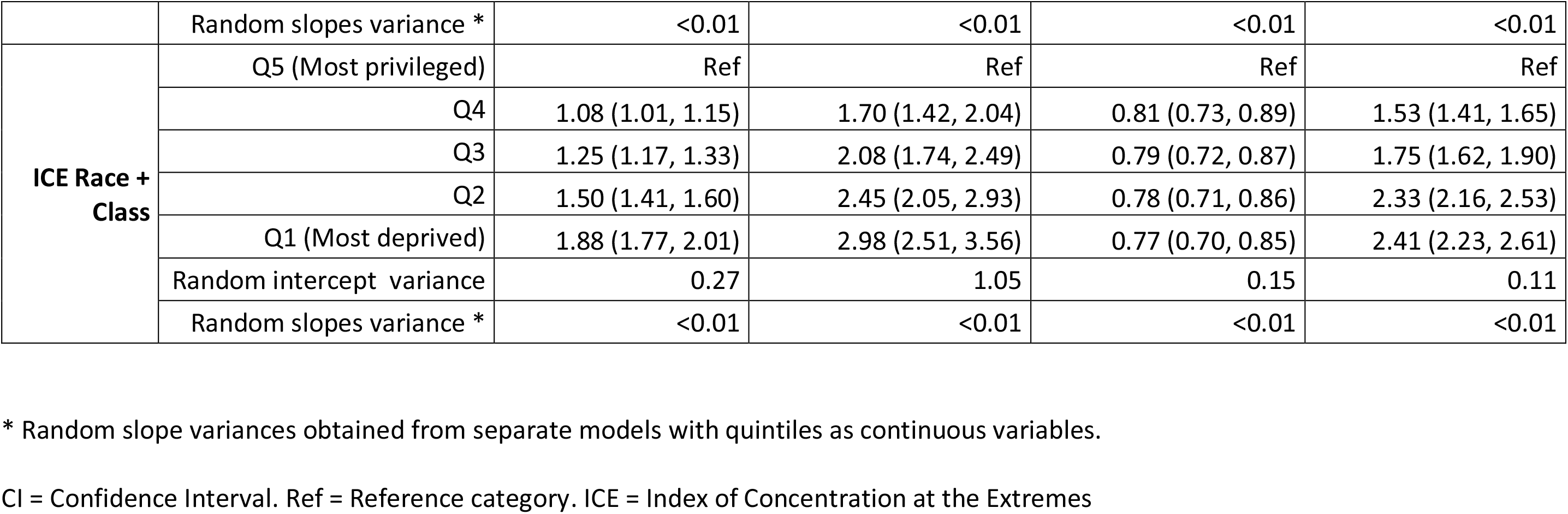
Cumulative Incidence Rate Ratios for Cases, Deaths and Tests, and Test Positivity Rate Ratios for quintiles of area-based socioeconomic measures, from multilevel Poisson models.

**Figure 1:**
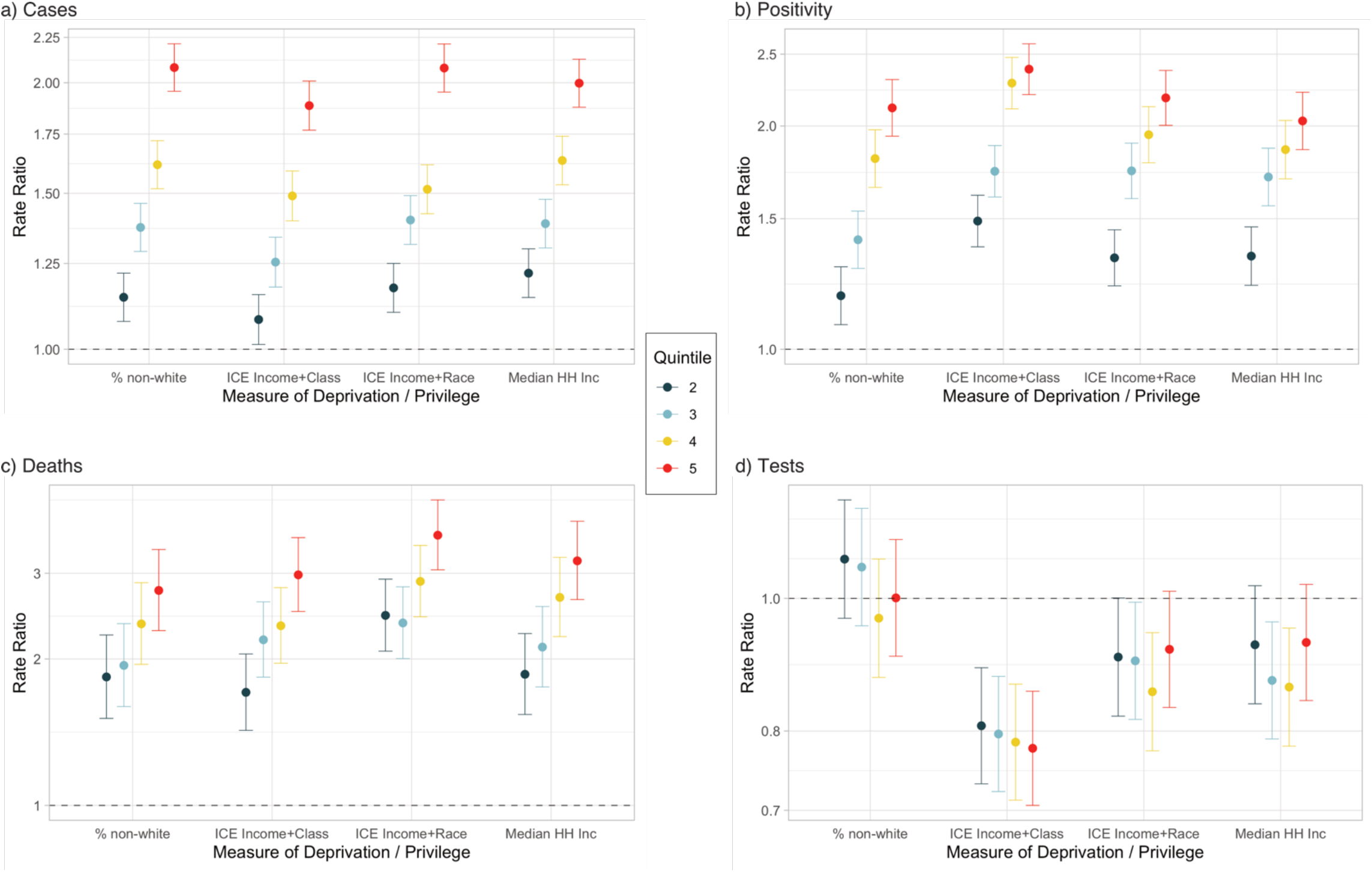
Rate ratios for COVID-19 a) cases, b) positivity, c) deaths, and d) tests from multilevel Poisson models for quintiles of Median Household Income, Percentage of non-white population, Index of Concentration at the Extremes for Race and Income, and Race and Working Class. The most privileged quintile is the referent category for each measure

Models with random slopes for quintiles as continuous variables suggested relatively little variance in the socioeconomic inequities between cities. For all measures and outcomes, the random slopes variance was 0.01 or less **(Table 2)**.

## DISCUSSION

In this study, we investigated neighborhood-level inequities in COVID-19 across cities in the US and Canada. While previous research has separately looked at how some counties have been disproportionately affected (Adhikari et al., 2020; Millett et al., 2020), and how areas within *specific* counties/cities have been disproportionately affected (Benitez et al., 2020; Chen and Krieger, 2020; Cordes and Castro, 2020; Lieberman-Cribbin et al., 2020), we are able to comprehensively report inequities across a large number of cities. We have compiled a dataset of neighborhood-level outcomes in a large number of urban regions, and have made this publicly available for researchers. We also present city-specific rates and trends in an accompanying website. In this dataset, we found considerable variation within cities. For example, the difference between the urban regions with the highest (Miami-Dade County: 5972 per 100,000) and second highest number of infections (Maricopa County: 3166 per 100,000) was 2800 per 100,000, but the difference between the most privileged quintile in Miami-Dade and the most deprived quintile in Maricopa burden (in terms of ICE race + class) was just 730 per 100,000.

Across cities, we found highly consistent and strong social gradients in rates of COVID-19 diagnosed cases, deaths and test positivity. However, testing rates did not follow the same social gradients, suggesting that testing efforts may not have been appropriately prioritized to address the most impacted populations and neighborhoods. Consequently, cases were likely underestimated in the most deprived neighborhoods. We found greater inequities in mortality compared to cases, which may reflect the fact that COVID-19 deaths are more likely to be ascertained than cases, and/or the underlying population characteristics that increase risk of death after infection (e.g. comorbidities, lack of health insurance).

Our findings are also broadly consistent with racial capitalism as a fundamental cause of COVID-19, with joint measures of racial and income or occupational segregation highlighting strong gradients of inequities. It is important to note that these are not proposed as measures of racial capitalism, per se. As proposed by Link and Phelan (Link and Phelan, 1995), and applied by Pirtle and McClure to racial capitalism (McClure et al., 2020; Pirtle, 2020), fundamental causes “…embody access to important resources, affect multiple disease outcomes through multiple mechanisms, and consequently maintain an association with disease even when intervening mechanisms change.” As a fundamental cause, racial capitalism is a structure that can be interrogated and investigated by different research questions and methods in different domains. Thus our study fits into this theoretical framework in conjunction with other evidence such as: racial occupational segregation such that Black and Hispanic / Latino workers are over-represented in frontline, high-risk and essential jobs; narratives of nativity in driving such occupational segregation (Hawkins, 2020; Queneau, 2009; Selden and Berdahl, 2020); racial and ethnic disparities in workplace outbreaks of COVID-19 (Bui, 2020; Waltenburg et al., 2020); both occupational and racial disparities in COVID-19 cases and deaths (Office for National Statistics, 2020; Peel Health Surveillance, 2020); and inequities in ability to work from home and socially distance after implementation of non-pharmaceutical interventions (Weill et al., 2020). Furthermore, racial capitalism in housing (e.g. redlining, predatory lending, evictions) can lead to spatial COVID-19 inequities directly due to spatial residential segregation and isolation (Dorries et al., 2019; Pulido, 2016), and indirectly through higher risk of household transmission stemming from poor and overcrowded housing conditions (Acevedo-Garcia, 2000). Our findings of consistent COVID-19 inequities across a range of cities, and two countries, also demonstrate how social processes embedded in fundamental causes can distribute risks of exposure in similar ways across populations.

Our study has a number of limitations, largely due to data quality issues affecting COVID-19 data generally. For case data, only a subset of all infections in the population are detected, subject to variations in testing criteria and access over time and between jurisdictions. This may affect comparison of case burden across cities, but it is unlikely to lead to overestimation of inequities within cities (Pitzer et al., 2020). In fact, within the subset of regions where we had test data, because test positivity was higher in deprived neighborhoods, and because a higher fraction of tests being positive suggests under-ascertainment of infections, our study likely underestimates inequities in infection risk. Deaths attributed to COVID-19 are also generally less than the surge in excess deaths that are observed, which may partly be due to under-ascertainment of COVID-19 as a cause of death (Chen et al., 2020). In presenting cumulative incidence proportions, we bias results such that regions with earlier onset of community transmission (e.g. New York City) have higher burdens compared to those with later onset. Ideally, the denominator would be person-time exposed from the time that some threshold of community transmission was passed. However, given the complex transmission dynamics, multiple introductions of COVID-19 into communities, and changing non-pharmaceutical interventions, it is difficult to do so consistently. Thus, while between-city differences should be interpreted with this caveat, we expect our within-city social gradients to still be indicative of persistent inequities. Next, different jurisdictions may report testing data differently as number of people tested, or number total tests administered. Finally, cases from community transmission versus in institutional settings (e.g. prisons, long-term care centers) are not consistently reported in an disaggregated matter. This introduces the potential for numbers being highly inflated in neighborhoods that host an affected institution. This is especially true for death rates since the infection fatality rate is high among the aged population in settings like long-term care centers.

Despite these limitations, our findings are consistent with reported racial/ethnic and socioeconomic inequities in COVID-19. When considered within the theoretical framework of racial capitalism, we believe that interventions that take into account the mutual forces of racism and capitalism will be effective in reducing both overall COVID-19 cases and inequities. For example, workplace-based interventions can be successful in reducing racial/ethnic and socioeconomic inequities (Almendrala, 2020). Our results also point to the importance of monitoring socio-economic inequities in addition to aggregate numbers, for both outcomes and testing efforts. This should be done through explicit measures of inequality and segregation, rather than just presenting spatial heterogeneity in geocoded cases through maps.

## Data Availability

Data, code, and additional visualizations available at: https://ssaha.shinyapps.io/COVID_cities_inequities

## SUPPLEMENTARY TABLES

**Supplementary Table 1.**
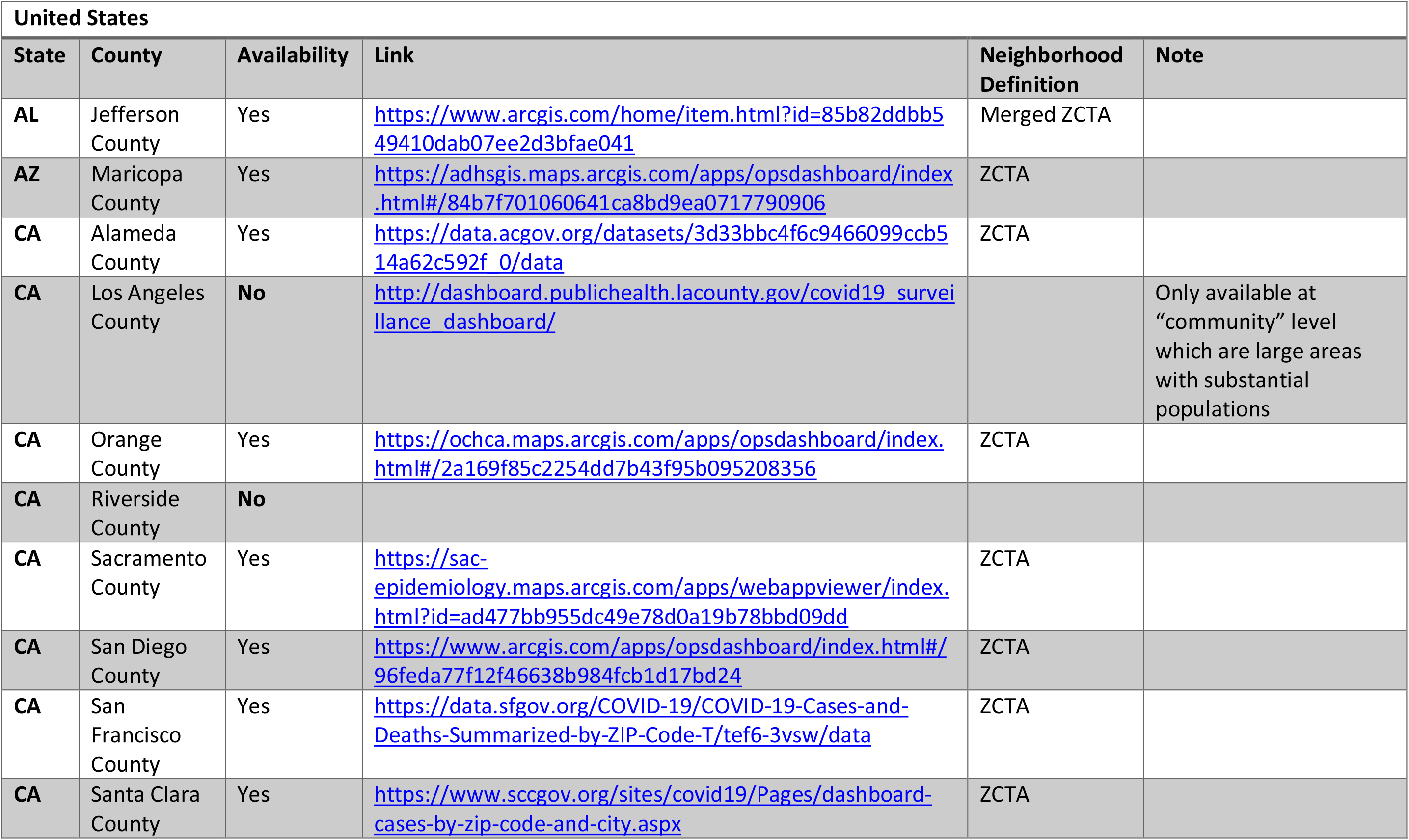
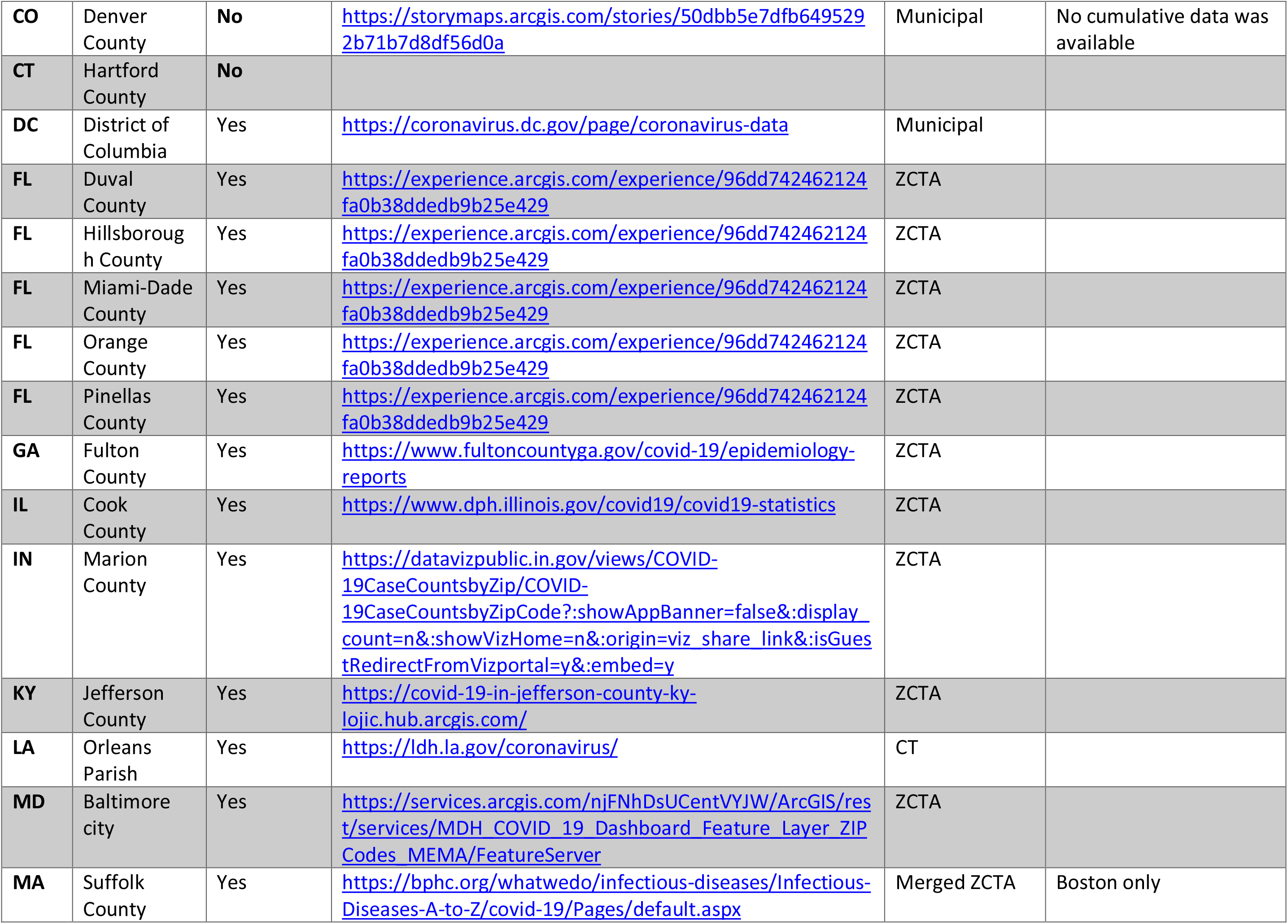
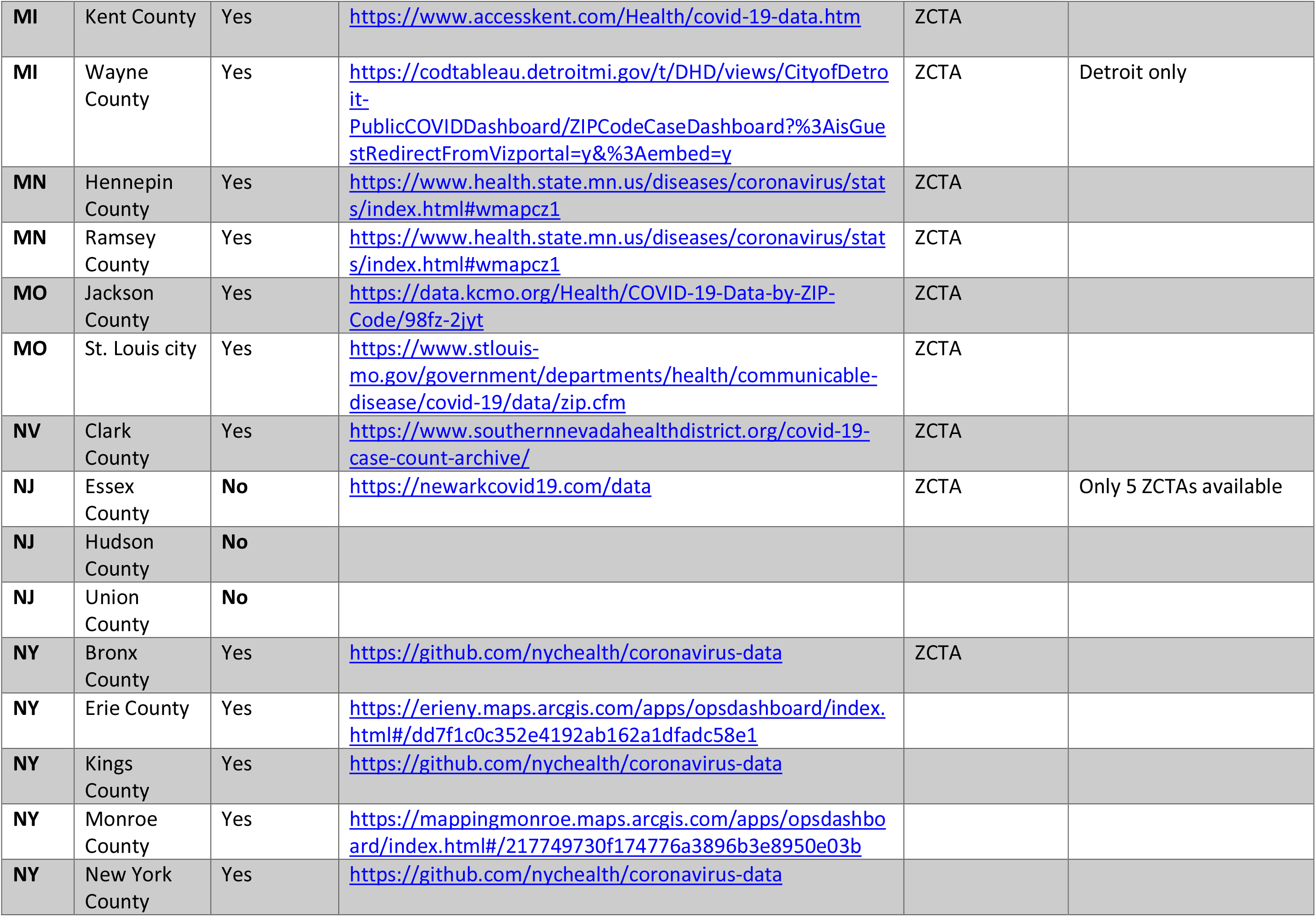
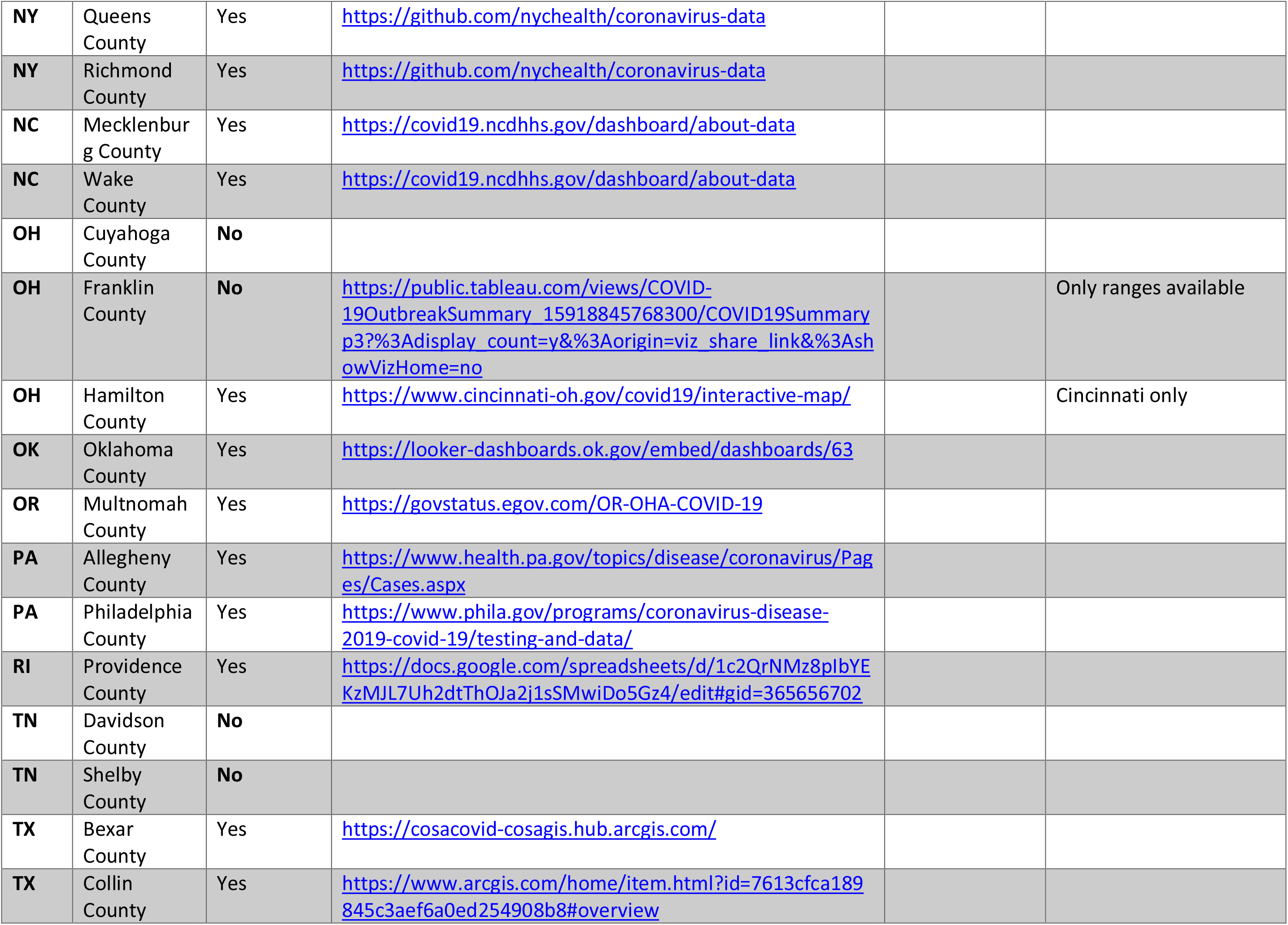
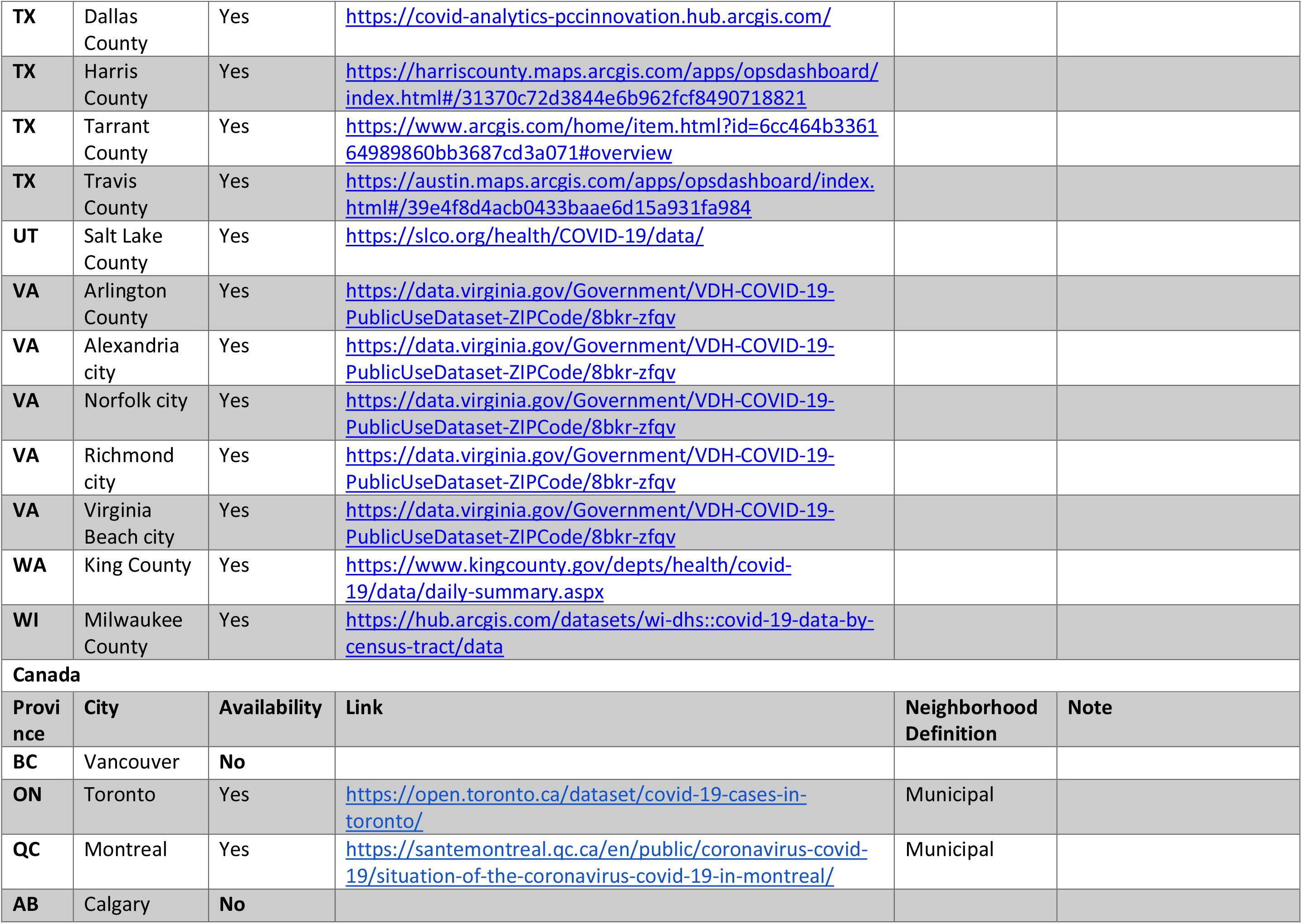
Data Availability and Sources.

| United States |  |  |  |  |  |
| --- | --- | --- | --- | --- | --- |
| State | County | Availability | Link | Neighborhood Definition | Note |
| AL | Jefferson County | Yes | https://www.arcgis.com/home/item.html?id=85b82ddb549410dab07ee2d3bfae041 | Merged ZCTA |  |
| AZ | Maricopa County | Yes | https://adhsgis.maps.arcgis.com/apps/opsdashboard/index.html#/84b7f701060641ca8bd9ea0717790906 | ZCTA |  |
| CA | Alameda County | Yes | https://data.acgov.org/datasets/3d33bbc4f6c9466099ccb514a62c592f_0/data | ZCTA |  |
| CA | Los Angeles County | No | http://dashboard.publichealth.lacounty.gov/covid19_surveillance_dashboard/ |  | Only available at “community” level which are large areas with substantial populations |
| CA | Orange County | Yes | https://ochca.maps.arcgis.com/apps/opsdashboard/index.html#/2a169f85c2254dd7b43f95b095208356 | ZCTA |  |
| CA | Riverside County | No |  |  |  |
| CA | Sacramento County | Yes | https://sac-epidemiology.maps.arcgis.com/apps/webappviewer/index.html?id=ad477bb955dc49e78d0a19b78bbd09dd | ZCTA |  |
| CA | San Diego County | Yes | https://www.arcgis.com/apps/opsdashboard/index.html#/96feda77f12f46638b984fcb1d17bd24 | ZCTA |  |
| CA | San Francisco County | Yes | https://data.sfgov.org/COVID-19/COVID-19-Cases-and-Deaths-Summarized-by-ZIP-Code-T/tef6-3vsw/data | ZCTA |  |
| CA | Santa Clara County | Yes | https://www.sccgov.org/sites/covid19/Pages/dashboard-cases-by-zip-code-and-city.aspx | ZCTA |  |
| **CO** | Denver County | No | https://storymaps.arcgis.com/stories/50dbb5e7dfb6495292b71b7d8df56d0a | Municipal | No cumulative data was available |
| **CT** | Hartford County | No |  |  |  |
| **DC** | District of Columbia | Yes | https://coronavirus.dc.gov/page/coronavirus-data | Municipal |  |
| **FL** | Duval County | Yes | https://experience.arcgis.com/experience/96dd742462124fa0b38ddedb9b25e429 | ZCTA |  |
| **FL** | Hillsborough County | Yes | https://experience.arcgis.com/experience/96dd742462124fa0b38ddedb9b25e429 | ZCTA |  |
| **FL** | Miami-Dade County | Yes | https://experience.arcgis.com/experience/96dd742462124fa0b38ddedb9b25e429 | ZCTA |  |
| **FL** | Orange County | Yes | https://experience.arcgis.com/experience/96dd742462124fa0b38ddedb9b25e429 | ZCTA |  |
| **FL** | Pinellas County | Yes | https://experience.arcgis.com/experience/96dd742462124fa0b38ddedb9b25e429 | ZCTA |  |
| **GA** | Fulton County | Yes | https://www.fultoncountyga.gov/covid-19/epidemiology-reports | ZCTA |  |
| **IL** | Cook County | Yes | https://www.dph.illinois.gov/covid19/covid19-statistics | ZCTA |  |
| **IN** | Marion County | Yes | https://datavizpublic.in.gov/views/COVID-19CaseCountsbyZip/COVID-19CaseCountsbyZipCode?:showAppBanner=false&:displaycount=n&:showVizHome=n&:origin=viz_share_link&:isGuestRedirectFromVizportal=y&:embed=y | ZCTA |  |
| **KY** | Jefferson County | Yes | https://covid-19-in-jefferson-county-ky-lojic.hub.arcgis.com/ | ZCTA |  |
| **LA** | Orleans Parish | Yes | https://ldh.la.gov/coronavirus/ | CT |  |
| **MD** | Baltimore city | Yes | https://services.arcgis.com/njFNhDsUCentVYJW/ArcGIS/rest/services/MDH_COVID_19_Dashboard_Feature_Layer_ZIP_Codes_MEMA/FeatureServer | ZCTA |  |
| **MA** | Suffolk County | Yes | https://bphc.org/whatwedo/infectious-diseases/Infectious-Diseases-A-to-Z/covid-19/Pages/default.aspx | Merged ZCTA | Boston only |
| **MI** | Kent County | Yes | https://www.accesskent.com/Health/covid-19-data.htm | ZCTA |  |
| **MI** | Wayne County | Yes | https://codtableau.detroitmi.gov/t/DHD/views/CityofDetroit-PublicCOVIDDashboard/ZIPCodeCaseDashboard?%3AisGuestRedirectFromVizportal=y&%3Aembed=y | ZCTA | Detroit only |
| **MN** | Hennepin County | Yes | https://www.health.state.mn.us/diseases/coronavirus/stats/index.html#wmapcz1 | ZCTA |  |
| **MN** | Ramsey County | Yes | https://www.health.state.mn.us/diseases/coronavirus/stats/index.html#wmapcz1 | ZCTA |  |
| **MO** | Jackson County | Yes | https://data.kcmo.org/Health/COVID-19-Data-by-ZIP-Code/98fz-2jyt | ZCTA |  |
| **MO** | St. Louis city | Yes | https://www.stlouis-mo.gov/government/departments/health/communicable-disease/covid-19/data/zip.cfm | ZCTA |  |
| **NV** | Clark County | Yes | https://www.southernnevadahealthdistrict.org/covid-19-case-count-archive/ | ZCTA |  |
| **NJ** | Essex County | **No** | https://newarkcovid19.com/data | ZCTA | Only 5 ZCTAs available |
| **NJ** | Hudson County | **No** |  |  |  |
| **NJ** | Union County | **No** |  |  |  |
| **NY** | Bronx County | Yes | https://github.com/nychealth/coronavirus-data | ZCTA |  |
| **NY** | Erie County | Yes | https://erieny.maps.arcgis.com/apps/opsdashboard/index.html#/dd7f1c0c352e4192ab162a1dfadc58e1 |  |  |
| **NY** | Kings County | Yes | https://github.com/nychealth/coronavirus-data |  |  |
| **NY** | Monroe County | Yes | https://mappingmonroe.maps.arcgis.com/apps/opsdashboard/index.html#/217749730f174776a3896b3e8950e03b |  |  |
| **NY** | New York County | Yes | https://github.com/nychealth/coronavirus-data |  |  |
| **NY** | Queens County | Yes | https://github.com/nychealth/coronavirus-data |  |  |
| **NY** | Richmond County | Yes | https://github.com/nychealth/coronavirus-data |  |  |
| **NC** | Mecklenburg County | Yes | https://covid19.ncdhhs.gov/dashboard/about-data |  |  |
| **NC** | Wake County | Yes | https://covid19.ncdhhs.gov/dashboard/about-data |  |  |
| **OH** | Cuyahoga County | **No** |  |  |  |
| **OH** | Franklin County | **No** | https://public.tableau.com/views/COVID-19OutbreakSummary_15918845768300/COVID19Summary_p3?%3Adisplay_count=y&%3Aorigin=viz_share_link&%3AshowVizHome=no |  | Only ranges available |
| **OH** | Hamilton County | Yes | https://www.cincinnati-oh.gov/covid19/interactive-map/ |  | Cincinnati only |
| **OK** | Oklahoma County | Yes | https://looker-dashboards.ok.gov/embed/dashboards/63 |  |  |
| **OR** | Multnomah County | Yes | https://govstatus.egov.com/OR-OHA-COVID-19 |  |  |
| **PA** | Allegheny County | Yes | https://www.health.pa.gov/topics/disease/coronavirus/Pages/Cases.aspx |  |  |
| **PA** | Philadelphia County | Yes | https://www.phila.gov/programs/coronavirus-disease-2019-covid-19/testing-and-data/ |  |  |
| **RI** | Providence County | Yes | https://docs.google.com/spreadsheets/d/1c2QrNMz8pIbYEKzMJL7Uh2dtThOJa2j1sSMwiDo5Gz4/edit#gid=365656702 |  |  |
| **TN** | Davidson County | **No** |  |  |  |
| **TN** | Shelby County | **No** |  |  |  |
| **TX** | Bexar County | Yes | https://cosacovid-cosagis.hub.arcgis.com/ |  |  |
| **TX** | Collin County | Yes | https://www.arcgis.com/home/item.html?id=7613cfca189845c3aef6a0ed254908b8#overview |  |  |
| **TX** | Dallas County | Yes | https://covid-analytics-pccinnovation.hub.arcgis.com/ |  |  |
| **TX** | Harris County | Yes | https://harriscounty.maps.arcgis.com/apps/opsdashboard/index.html#/31370c72d3844e6b962fcf8490718821 |  |  |
| **TX** | Tarrant County | Yes | https://www.arcgis.com/home/item.html?id=6cc464b336164989860bb3687cd3a071#overview |  |  |
| **TX** | Travis County | Yes | https://austin.maps.arcgis.com/apps/opsdashboard/index.html#/39e4f8d4acb0433baae6d15a931fa984 |  |  |
| **UT** | Salt Lake County | Yes | https://slco.org/health/COVID-19/data/ |  |  |
| **VA** | Arlington County | Yes | https://data.virginia.gov/Government/VDH-COVID-19-PublicUseDataset-ZIPCode/8bkr-zfqv |  |  |
| **VA** | Alexandria city | Yes | https://data.virginia.gov/Government/VDH-COVID-19-PublicUseDataset-ZIPCode/8bkr-zfqv |  |  |
| **VA** | Norfolk city | Yes | https://data.virginia.gov/Government/VDH-COVID-19-PublicUseDataset-ZIPCode/8bkr-zfqv |  |  |
| **VA** | Richmond city | Yes | https://data.virginia.gov/Government/VDH-COVID-19-PublicUseDataset-ZIPCode/8bkr-zfqv |  |  |
| **VA** | Virginia Beach city | Yes | https://data.virginia.gov/Government/VDH-COVID-19-PublicUseDataset-ZIPCode/8bkr-zfqv |  |  |
| **WA** | King County | Yes | https://www.kingcounty.gov/depts/health/covid-19/data/daily-summary.aspx |  |  |
| **WI** | Milwaukee County | Yes | https://hub.arcgis.com/datasets/wi-dhs::covid-19-data-by-census-tract/data |  |  |
| **Canada** |  |  |  |  |  |
| **Province** | **City** | **Availability** | **Link** | **Neighborhood Definition** | **Note** |
| **BC** | Vancouver | **No** |  |  |  |
| **ON** | Toronto | Yes | https://open.toronto.ca/dataset/covid-19-cases-in-toronto/ | Municipal |  |
| **QC** | Montreal | Yes | https://santemontreal.qc.ca/en/public/coronavirus-covid-19/situation-of-the-coronavirus-covid-19-in-montreal/ | Municipal |  |
| **AB** | Calgary | **No** |  |  |  |

**Supplementary Table 2.** Cumulative Test, Incidence and Death Proportions by Urban County / City

| Urban Area | Cases | Tests | Deaths | Population | Cumulative Test Rate per 100 (95% CI) | Cumulative Incidence Rate per 100,000 (95% CI) | Death Rate per 100,000 (95% CI) | Positivity Rate |
| --- | --- | --- | --- | --- | --- | --- | --- | --- |
| Maricopa County, AZ | 136994 | - | - | 4325769 | - | 3166.93<br>(3150.18, 3183.74) | - | - |
| Alameda County, CA | 19295 | 433921 | - | 1643825 | 26.40<br>(26.32, 26.45) | 1173.79<br>(1157.28, 1190.47) | - | 4.45<br>(4.38, 4.51) |
| Orange and San Diego Counties, CA | 88237 | - | 1035 | 6553139 | - | 1346.48<br>(1337.61, 1355.40) | 15.79<br>(14.85, 16.79) | - |
| Sacramento County, CA | 18780 | - | - | 1508357 | - | 1245.06<br>(1227.32, 1263.00) | - | - |
| San Francisco and Santa Clara Counties, CA | 28509 | - | 89 | 2768625 | - | 1029.72<br>(1017.80, 1041.74) | 3.21<br>(2.58, 3.96) | - |
| Duval County, FL | 28999 | - | - | 967181 | - | 2998.3<br>(2963.89, 3033.01) | - | - |
| Orange County, FL | 38329 | - | - | 1415274 | - | 2708.24<br>(2681.19, 2735.49) | - | - |
| Miami-Dade County, FL | 162300 | - | - | 2715526 | - | 5976.74<br>(5947.70, 6005.89) | - | - |
| Hillsborough and Pinellas Counties, FL | 60753 | - | - | 2420687 | - | 2509.74<br>(2489.82, 2529.78) | - | - |
| Fulton County, GA | 24049 | - | - | 1017566 | - | 2363.38<br>(2333.61, 2393.45) | - | - |
| Cook County, IL | 138852 | 1845575 | - | 5532272 | 33.36<br>(33.31, 33.41) | 2509.85<br>(2496.67, 2523.09) | - | 7.52<br>(7.48, 7.56) |
| **Marion County, IN** | 17860 | - | - | 1001224 | - | 1783.82<br>(1757.75, 1810.17) | - | - |
| **Jefferson County, KY** | 12838 | - | - | 765575 | - | 1676.91<br>(1648.03, 1706.17) | - | - |
| **Baltimore County, MD** | 23336 | - | - | 1027396 | - | 2271.37<br>(2242.32, 2300.71) | - | - |
| **Ramsey and Hennepin**<br>**Counties, MN** | 37322 | - | - | 1955088 | - | 1908.97<br>(1889.65, 1928.43) | - | - |
| **Kansas City, MO** | 6650 | 65713 | - | 340595 | 19.29<br>(19.15, 19.44) | 1952.46<br>(1905.82, 1999.97) | - | 10.12<br>(9.88, 10.37) |
| **St Louis County, MO** | 6442 | - | - | 312358 | - | 2062.38<br>(2012.32, 2113.37) | - | - |
| **Clark County, NV** | 60940 | - | - | 2141583 | - | 2845.56<br>(2823.01, 2868.24) | - | - |
| **Erie County, NY** | 10135 | - | - | 913920 | - | 1108.96<br>(1087.47, 1130.76) | - | - |
| **Monroe County, NY** | 5436 | - | - | 743595 | - | 731.04<br>(711.74, 750.74) | - | - |
| **Wake County, NC** | 20524 | - | 264 | 1249624 | - | 1642.41<br>(1620.02, 1665.04) | 21.13<br>(18.65, 23.83) | - |
| **Mecklenburg County,**<br>**NC** | 27431 | - | 346 | 1107116 | - | 2477.7<br>(2448.46, 2507.2) | 31.25<br>(28.05, 34.73) | - |
| **Oklahoma County, OK** | 15656 | - | 175 | 841965 | - | 1859.46<br>(1830.45, 1888.82) | 20.78<br>(17.82, 24.1) | - |
| **Multnomah County,**<br>**OR** | 2639 | - | - | 992484 | - | 265.9<br>(255.85, 276.24) | - | - |
| **Philadelphia County,**<br>**PA** | 35205 | 355948 | 1761 | 1575729 | 22.59<br>(22.52, 22.66) | 2234.2<br>(2210.93, 2257.67) | 111.76<br>(106.6, 117.1) | 9.89<br>(9.79, 9.99) |
| **Alleghany County, PA** | 11846 | 178215 | - | 1408425 | 12.65<br>(12.60, 12.71) | 841.08<br>(826.00, 856.37) | - | 6.65<br>(6.53, 6.77) |
| **Providence County, RI** | 17574 | - | - | 667425 | - | 2633.11<br>(2594.32, 2672.33) | - | - |
| **Bexar County, TX** | 40861 | - | - | 1926908 | - | 2120.55<br>(2100.04, 2141.21) | - | - |
| **Collin, Dallas, and Tarrant Counties, TX** | 121746 | - | - | 6316898 | - | 1927.31<br>(1916.5, 1938.16) | - | - |
| **Harris County, TX** | 116927 | - | 1575 | 4593398 | - | 2545.54<br>(2530.98, 2560.18) | 34.29<br>(32.62, 36.02) | - |
| **Travis County, TX** | 27010 | - | - | 1202776 | - | 2245.64<br>(2218.94, 2272.58) | - | - |
| **Washington, D.C. and Arlington and Alexandria Counties, VA** | 22640 | 410034 | - | 1129965 | 36.29<br>(36.18, 36.40) | 2003.60<br>(1977.59, 2029.87) | - | 5.52<br>(5.45, 5.59) |
| **Richmond County, VA** | 7712 | 108500 | - | 379033 | 28.63<br>(28.46, 28.80) | 2034.65<br>(1989.49, 2080.58) | - | 7.11<br>(6.95, 7.27) |
| **Virginia and Norfolk Counties, VA** | 10550 | 153256 | - | 695654 | 22.03<br>(21.92, 22.14) | 1516.56<br>(1487.76, 1545.78) | - | 6.88<br>(6.75, 7.02) |
| **King County, WA** | 20132 | 378045 | - | 2163584 | 17.47<br>(17.42, 17.53) | 930.49<br>(917.68, 943.44) | - | 5.33<br>(5.25, 5.4) |
| **Kent County, MI** | 8108 | - | - | 642064 | - | 1262.80<br>(1235.46, 1290.59) | - | - |
| **City of Detroit, MI** | 13037 | - | 1439 | 701564 | - | 1858.28<br>(1826.51, 1890.45) | 205.11<br>(194.65, 215.99) | - |
| **City of Cincinnati, OH** | 5101 | - | 101 | 473213 | - | 1077.95<br>(1048.57, 1107.95) | 21.34<br>(17.38, 25.93) | - |
| **Salt Lake County, UT** | 24347 | - | - | 1120013 | - | 2173.81<br>(2146.59, 2201.29) | - | - |
| **Milwaukee County, WI** | 15245 | 143797 | - | 914044 | 15.73<br>(15.65, 15.81) | 1667.86<br>(1641.49, 1694.55) | - | 10.6<br>(10.43, 10.77) |
| **New Orleans Parish, LA** | 10305 | 168238 | - | 389648 | 43.18<br>(42.97, 43.38) | 2644.69<br>(2593.88, 2696.26) | - | 6.13<br>(6.01, 6.24) |
| **Jefferson County, AL** | 18037 | - | - | 661217 | - | 2727.85<br>(2688.18, 2767.95) | - |  |
| **New York City, NY** | 227344 | 2171108 | 19081 | 8443637 | 25.71<br>(25.68, 25.75) | 2692.49<br>(2681.43, 2703.58) | 225.98<br>(222.79, 229.21) | 10.47<br>(10.43, 10.51) |
| **City of Boston, MA** | 15846 | - | - | 673934 | - | 2351.27<br>(2314.8, 2388.17) | - | - |
| **City of Toronto, ON, Canada** | 15783 | - | 1133 | 2731571 | - | 577.80<br>(568.82, 586.89) | 41.48<br>(39.1, 43.97) | - |
| **City of Montréal, QC, Canada** | 29811 | - | 3421 | 1941700 | - | 1535.30<br>(1517.92, 1552.83) | 176.19<br>(170.33, 182.19) | - |

**Supplementary Table 3:**
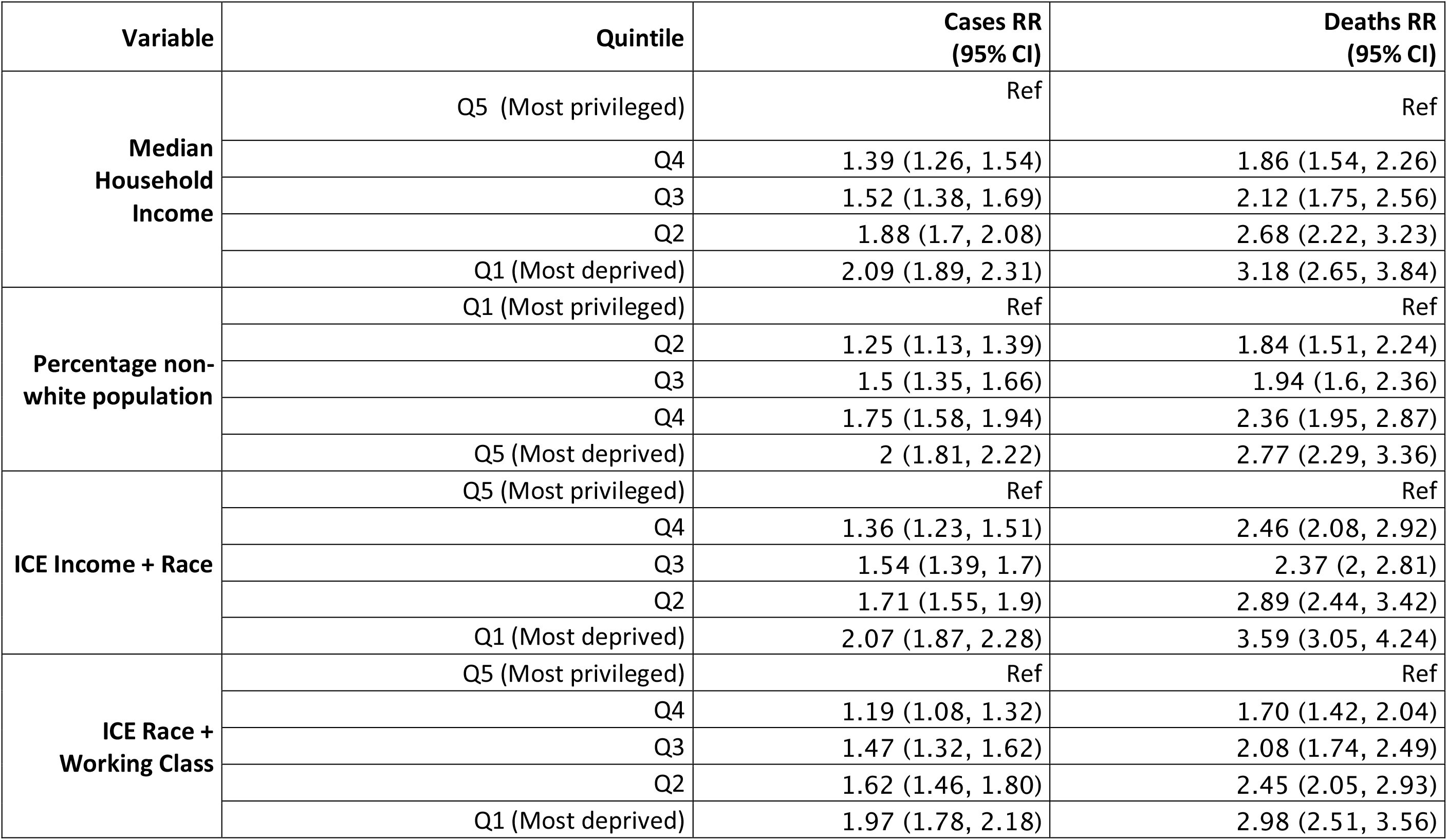
Cumulative Incidence Rate Ratios for cases and deaths for quintiles of area-based socioeconomic measures, from multilevel Poisson models, among urban regions that have data for both outcomes

**Supplementary Table 4:** Cumulative Incidence Rate Ratios for cases and tests for quintiles of area-based socioeconomic measures, from multilevel Poisson models, among urban regions that have data for both outcomes

| Variable | Quintile | Cases RR<br>(95% CI) | Tests RR<br>(95% CI) |
| --- | --- | --- | --- |
| **Median Household Income** | Q5 (Most privileged) | Ref | Ref |
|  | Q4 | 1.25 (1.13, 1.38) | 0.92 (0.84, 1.02) |
|  | Q3 | 1.49 (1.36, 1.65) | 0.87 (0.79, 0.96) |
|  | Q2 | 1.66 (1.51, 1.84) | 0.86 (0.78, 0.95) |
|  | Q1 (Most deprived) | 1.93 (1.75, 2.12) | 0.93 (0.84, 1.02) |
| **Percentage non-white population** | Q1 (Most privileged) | Ref | Ref |
|  | Q2 | 1.24 (1.12, 1.36) | 1.07 (0.97, 1.18) |
|  | Q3 | 1.44 (1.31, 1.58) | 1.05 (0.95, 1.16) |
|  | Q4 | 1.74 (1.58, 1.92) | 0.97 (0.88, 1.07) |
|  | Q5 (Most deprived) | 2.09 (1.91, 2.3) | 1.00 (0.91, 1.1) |
| **ICE Income + Race** | Q5 (Most privileged) | Ref | Ref |
|  | Q4 | 1.21 (1.1, 1.34) | 0.91 (0.82, 1) |
|  | Q3 | 1.56 (1.41, 1.71) | 0.9 (0.82, 0.99) |
|  | Q2 | 1.69 (1.53, 1.86) | 0.85 (0.77, 0.94) |
|  | Q1 (Most deprived) | 2.03 (1.85, 2.23) | 0.92 (0.83, 1.01) |
| **ICE Race + Working Class** | Q5 (Most privileged) | Ref | Ref |
|  | Q4 | 1.26 (1.15, 1.39) | 0.81 (0.73, 0.89) |
|  | Q3 | 1.40 (1.27, 1.54) | 0.79 (0.72, 0.87) |
|  | Q2 | 1.83 (1.66, 2.02) | 0.78 (0.71, 0.87) |
|  | Q1 (Most deprived) | 1.93 (1.76, 2.12) | 0.77 (0.70, 0.85) |

